# Synapse loss in Progressive Supranuclear Palsy *post-mortem* reflects clinical and pathological disease severity and ^11^C-UCB-J PET *in vivo*

**DOI:** 10.64898/2026.06.02.26354325

**Authors:** George Nolan, Negin Holland, Shangqing W. Yang, Ginevra Matilde Dall’Ò, Qiyuan Chen, Kieren Allinson, George Savulich, Kristy Halliday, Michelle Naessens, Young T. Hong, Tim D. Fryer, Franklin I. Aigbirhio, Maura Malpetti, Sanne Simone Kaalund, John T. O’Brien, András Lakatos, James B. Rowe, Annelies Quaegebeur

## Abstract

Synapse loss is an early feature of neurodegeneration and may provide sensitive biomarkers for experimental medicine. Positron emission tomography (PET) with the synaptic vesicle glycoprotein 2A radioligand [¹¹C]UCB-J shows widespread signal reduction across dementias. However, it remains unclear which aspects of synaptic integrity [¹¹C]UCB-J PET measures.

We developed a histological-imaging pipeline to quantify structurally intact synapses in post-mortem brain tissue. We applied it to six donors with the tauopathy progressive supranuclear palsy (PSP) who had ante-mortem [¹¹C]UCB-J-PET, alongside six controls across 11 brain regions.

Synapse loss in PSP was widespread but region-specific across cortical, subcortical, and brainstem regions. Greater synapse loss was associated with higher tau burden and pathology, and cortical synaptic density correlated with ante-mortem cognition. Post-mortem synaptic density correlated with *in vivo* [¹¹C]UCB-J-PET signal.

This study provides validation of SV2A PET as a biomarker of synaptic density and supports integration of imaging with histopathology in neurodegenerative disease research.

## Background

Experimental and clinical studies identify synapse loss as an early, shared event across neurodegenerative diseases, including Alzheimer’s disease (AD) and frontotemporal lobar degeneration (FTLD) syndromes, such as the primary tauopathy progressive supranuclear palsy (PSP) [1,2]. In many experimental models, including tauopathies, synaptic degeneration precedes overt neuronal loss [3,4]. This vulnerability is driven by processes such as toxic misfolded proteins [5–8], neuroinflammation [9,10], oxidative stress [11] and altered glial activity [8,12,13]. Clinicopathological studies demonstrate that synaptic density, measured using single synaptic protein quantification or electron microscopy (EM), correlates strongly with cognitive decline in AD [14–18]. Together, these findings highlight synaptic loss as both a therapeutic target and an intermediate outcome marker of treatment response targeting upstream processes. Consequently, developing sensitive in vivo biomarkers to measure synaptic density across the brain capable of detecting early degeneration and tracking disease progression has become a major translational priority.

The PET radioligand [¹¹C]UCB-J enables in vivo synaptic imaging by binding to SV2A, a presynaptic vesicle protein widely expressed across the brain [19]. Regionally, SV2A abundance correlates with established presynaptic markers, such as synaptophysin [19,20], and autoradiography confirms that [³H]UCB-J binds specifically to SV2A and correlates to synaptotagmin and synaptophysin [21,22], supporting its use as an index of presynaptic integrity. However, recent single-synapse studies resolution imaging have questioned this interpretation. SV2A appears in only ∼25% of synapses in human temporal cortex and shows variable, non-linear expression across mouse brain regions [23]. Consequently, reduced [¹¹C]UCB-J binding may reflect not only synapse loss, but also changes in vesicle number or SV2A expression per vesicle [24]. One study found no significant difference in brain tissue [³H]UCB-J binding between controls and advanced AD cases [25]. These findings highlight uncertainty in how SV2A relates to true synapse number and underscore the need for systematic post-mortem validation linking [¹¹C]UCB-J-PET signal to structurally intact synapse density across brain regions.

Reduced [¹¹C]UCB-J binding has been interpreted as reflecting synaptic loss across multiple neurodegenerative conditions, including PSP, frontotemporal dementia (FTD), AD, corticobasal syndrome (CBS), Parkinson’s disease, Huntington’s disease, and Lewy body dementia [26–34] and is generally associated with greater disease severity. In PSP and amyloid-negative CBS, reduced frontal binding correlates with higher tau burden measured using [¹⁸F]AV-1451-PET, in subcortical afferents [27]. In presymptomatic genetic FTD, reductions are detectable years before symptom onset, supporting synapse loss as an early pathological event [35]. Longitudinally, reduced [¹¹C]UCB-J binding progresses rapidly in PSP (∼4% per year vs ∼1% per decade in controls) and correlates with clinical decline [28]. Together, these findings support the use of [¹¹C]UCB-J-PET to monitor disease progression and assess target engagement in clinical trials.

While neuropathological studies are essential for establishing the fidelity of [¹¹C]UCB-J PET as a measure of synaptic density, post-mortem synapse quantification remains technically challenging. Bulk protein analysis can demonstrate regional changes in synaptic proteins in FTD, PSP, and AD [36,37] but cannot distinguish intact synapses from degenerating terminals [38] or account for protein redistribution across compartments [39]. Proximity-based immunohistochemical methods, which identify apposed pre- and postsynaptic markers, allow more direct quantification of structurally intact synapses [40]. Given the uncertainty about how SV2A relates to true synapse number, correlating [¹¹C]UCB-J-PET signal with directly measured intact synapse density is likely more informative than comparison with SV2A levels alone.

PSP is a valuable model for studying PET-to-post-mortem relationships. It is a rapidly progressive 4-repeat tauopathy with severe motor and cognitive impairment and strong clinicopathological correlation [41–44]. Hallmark pathological features include neurofibrillary tangles, tufted astrocytes, and coiled bodies across affected cortical, subcortical, and brainstem regions [45]. PSP pathology can be reliably staged using validated tau distribution patterns [46,47], enabling systematic assessment of synaptic loss across disease progression.

This study aimed to: (i) develop an integrated histological and imaging pipeline to quantify post-mortem loss of structurally intact synapses in PSP; (ii) assess the relationship between tau burden and synaptic loss; (iii) examine associations between synapse density and ante-mortem cognitive performance; and (iv) determine whether post-mortem synapse density correlates with ante-mortem [¹¹C]UCB-J-PET binding across regions and individuals, directly testing its validity as a measure of synaptic density.

### Participant Characteristics

Six people with PSP from the Cambridge Centre for Parkinson-plus, who had undergone [^11^C]UCB-J PET imaging during life [26,28], donated their brain to the Cambridge Brain Bank. All six participants were clinically assessed ante-mortem with the Addenbrooke’s Cognitive Examination-revised [48] on average, 18 months prior to death. Six age-and gender-matched neurologically healthy brain donors with mild age-related neuropathology were selected as a control group. All tissue was obtained from the Cambridge Brain Bank under the Neuropathology Research in Dementia protocol (Research Ethics Committee reference 16/WA/0240). Participant characteristics and sampled brain regions are summarised in Table 1. Both ante-mortem and post-mortem correlations were adjusted for time between clinical observations and death, using proportions listed in Table 1.

**Table 1.** Demographic, clinical and pathological characteristics of control and PSP donors.

| Case | Age range at death (years) | Sex | Total symptom duration (months) | ACE-R score max 100 | Symptom duration at ACE-R (months) | ACE-R to death interval (months) | Proportion of symptom duration at ACE-R (%) | Symptom duration at PET (months) | PET to death interval (months) | Proportion of symptom duration at PET (%) | PSP tau pathology stage |
| --- | --- | --- | --- | --- | --- | --- | --- | --- | --- | --- | --- |
| 1 | 66-70 | F | 0 | NA | 0 | NA | 0 | 0 | 0 | 0 | 0 |
| 2 | 61-65 | F | 0 | NA | 0 | NA | 0 | 0 | 0 | 0 | 0 |
| 3 | 66-70 | F | 0 | NA | 0 | NA | 0 | 0 | 0 | 0 | 0 |
| 4 | 91-96 | M | 0 | NA | 0 | NA | 0 | 0 | 0 | 0 | 0 |
| 5 | 71-75 | M | 0 | NA | 0 | NA | 0 | 0 | 0 | 0 | 0 |
| 6 | 81-85 | M | 0 | NA | 0 | NA | 0 | 0 | 0 | 0 | 0 |
| 7 | 76-80 | M | 53.1 | 75 | 34.9 | 18.2 | 65.7 | 35.9 | 17.2 | 67.6 | 6 |
| 8 | 76-80 | F | 67.5 | 37 | 45.0 | 22.5 | 66.7 | 47.6 | 19.9 | 70.5 | 6 |
| 9 | 71-75 | F | 86.2 | 84 | 83.8 | 2.5 | 97.1 | 69.4 | 16.9 | 80.5 | 4 |
| 10 | 71-75 | M | 67.0 | 78 | 58.6 | 8.4 | 87.5 | 48.1 | 18.9 | 71.8 | 4 |
| 11 | 71-75 | M | 58.9 | 48 | 52.2 | 6.6 | 88.7 | 54.8 | 4.1 | 93.1 | 5 |
| 12 | 61-75 | F | 78.6 | 69 | 57.9 | 20.7 | 73.6 | 57.9 | 20.7 | 73.7 | 4 |

To validate *in vivo* [^11^C]UCB-J PET findings, non-displaceable binding potentials (BP_ND_) from this cohort were obtained using the neuroimaging methodology outlined [26,28]. We used [^11^C]UCB-J BP_ND_ values from the 11 Hammersmith Atlas brain regions outlined above (http://brain-development.org), corrected for the partial volume of cerebrospinal fluid in each region.

### Brain tissue preparation and synapse immunofluorescence

Frozen blocks from 11 selected brain regions from the right hemisphere were sampled by a neuropathologist and sectioned at 14µm sections for immunofluorescence. Regions were selected based on the Hammersmith Brain Atlas used for ante-mortem [^11^C]UCB-J PET scans [26] and their differential involvement by tau pathology: primary visual cortex, inferior frontal gyrus, precentral gyrus, middle temporal lobe, superior parietal lobule, anterior cingulate cortex, putamen, globus pallidus, thalamus, substantia nigra and tegmentum midbrain [46,47].

A four-plex immunofluorescence labelling protocol was optimised to visualise intact synapses with pre-synaptic Bassoon, post-synaptic Homer1, dendritic marker microtubule-associated protein 2 (MAP2) and nucleic marker DAPI. Synapses were quantified by the proximity and colocalisation of pre- and post-synaptic puncta. Candidate pre- and post-synaptic terminal markers were selected based on their spatial localisation, signal enrichment, and virtual sharpness, quantified by the full width at half maximum (FWHM) [49]. MAP2 staining delineated anatomical landmarks of regions of interest (ROI).

Sections were post-fixed in 4% PFA, permeabilised and blocked in PBS-saponin (0.1%) with 5% BSA at room temperature. Saponin was used due to its advantage over conventional detergents in preserving membranes and, by extension, membrane-bound antigens like synaptic markers [50]. Following overnight incubation with primary antibodies at 4°C, lipofuscin autofluorescence was quenched with TrueBlack, followed by 2.5 h secondary antibody incubation and DAPI counterstaining. Sections were mounted in Fluoromount-G.

### Tau immunohistochemistry and analysis

AT8 DAB-based immunohistochemistry was performed on corresponding formalin-fixed, paraffin-embedded (FFPE) brain sections from the left hemisphere. Tau pathology in PSP donors was staged by a neuropathologist, in accordance with Kovacs et al [46]. For regional tau burden quantification, whole slide images were acquired with Aperio AT2 brightfield whole-slide scanning (Leica, 20x) and analysed in QuPath. Cortical section ROIs were semi-automatically delineated into grey and white matter regions using pixel training classification of cerebellum training datasets. Subcortical and brainstem regions were manually delineated. Tau burden was quantified using a Gaussian-filtered pixel classifier based on size and colour. Tau area fraction was defined as the percentage of tau-positive area relative to total ROI tissue area.

### Synapse confocal acquisition

Trans-cortical tile scans were constructed to capture neocortical layers I-VI and subsequently divided into superficial, middle, deep cortical depth compartments. Subcortical ROIs were delineated using MAP2-defined landmarks on adjacent sections. Whole slide epifluorescence images were acquired on a Leica DMi8 microscope (0.25 NA, 10x objective) and Leica DFC365 FX camera and resulting merged images were delineated under guidance from a neuropathologist. Random fields were sampled within ROIs.

Confocal images were acquired on a Leica TCS SP8 (63×, 1.4 NA, HyD photon-counting detectors) using simultaneous 405/561 and 488/640 nm line acquisition. Images were collected at 1,024 × 1,024 resolution, 16-bit depth, pinhole 1 AU. Z-stacks used 0.8–1.0 µm steps (7–8 µm total) for cortex and 0.3 µm steps (4.5–6 µm total) for subcortical regions.

Tiles were exported to ImageJ, the optimal focal plane was selected by histogram peak position and analysed in CellProfiler. Cortical tile data were divided into 3 layers of depth (superficial, middle and deep, of equal depth). These subgroups were used for cortical depth-specific analysis.

### Synapse Density quantification

Images were analysed in CellProfiler v4.2.5. Synapses were defined by proximity of Bassoon and Homer1 immunoreactive signals representing the pre- and post-synaptic terminals, respectively. The IdentifyPrimaryObjects module was used and images were filtered by a Gaussian value of σ =1, thresholded via object size thresholding of 2-6 pixel diameter, Otsu adaptive intensity thresholding with an adaptive window of 6^2^ pixels and a manual threshold for both pre- and post-synaptic channels. The proximity of segmented objects was assessed using MeasureObjectsOverlaps and MeasureObjectNeighbour (maximum distance 20 pixels). Outputs were processed in Python (VSCode 3.11.9). Synapse identification used a bespoke Sydentify pipeline (https://github.com/QiyuanChen02/Sydentify), based on the coordinates of thresholded synapse labels and the proximity to the nearest opposite channel neighbour with a maximum distance for Bassoon and Homer1 immunoreactive synaptic boutons. An upper resolutional threshold (1 pixel = 180nm) was applied as a proximity threshold in view of the variability in the virtual distance of the synaptic cleft in immunofluorescence images [40,51] due to the planar orientation of antibody binding [52] or imaging artefacts (fluorescence halo).

### Statistical Analysis

Statistical analyses were performed in RStudio (v4.2.0) and Graphpad (v10.2.3). Post-mortem synaptic density was presented as mean synapses/µm² ± standard deviation (SD). Regional differences in synaptic density were assessed using one-way ANOVA. Homogeneity of variance was evaluated with the Brown–Forsythe test; where violated, Welch’s ANOVA was applied. Post-hoc regional comparisons were corrected for multiple testing using the Benjamini-Hochberg false discovery rate (FDR), and adjusted q-values are reported (Supplementary Table 1). Differences in synaptic density in function of cortical depth were analysed using linear mixed-effects models with group, region and cortical depth (superficial, middle, deep) as fixed effects and subject as a random effect. Synaptic density across PSP tau pathology stages was examined using two-way ANOVA or linear mixed-effects models, where appropriate, to test the interaction between region, disease stage and subject. Controls were assigned PSP tau pathology stage 0; PSP cases were tau pathology stages 4–6, with the single stage 5 case grouped with stage 6. Where sphericity or variance assumptions were not met, Geisser–Greenhouse correction was applied. Post-hoc pairwise comparisons (control vs stage 4 vs stage 5/6) were FDR-corrected. Both uncorrected and corrected q-values were reported (Table 2). For tau burden (area fraction), heterogeneity of variance across the PSP cohort was confirmed and Welch’s ANOVA applied, followed by regional post-hoc comparisons with FDR correction. Tau area fraction showed a lognormal distribution and was log-transformed for correlational analyses. In view of unequal variance, non-parametric Spearman rank correlations were used. Linear regression lines are shown for visualisation.

**Table 2.** Synaptic density in stage 4 (B) and 5/6 (C) tau pathology. Synaptic density reported as mean synapses/µm^2^ (± SD). *P*-values were derived from two-way ANOVA with Greenhouse–Geisser correction. Adjusted *p*-values were obtained from post hoc pairwise comparisons and corrected for multiple comparisons using false discovery rate (FDR) control. Post hoc comparisons were performed between Group A (controls), Group B (PSP tau pathology stage 4), and Group C (PSP tau pathology stage 5/6), with adjusted *p*-values reported for each group comparison.

| Region | (A) Control<br>synapses/$\mu\text{m}^2$ (SD) | (B) PSP tau<br>pathology stage 4<br>synapses/$\mu\text{m}^2$ (SD) | (C) PSP tau<br>pathology stage 5/6<br>synapses/$\mu\text{m}^2$ (SD) | A vs B<br>p value (p.adj) | A vs C<br>p value (p.adj) | B vs C<br>p value<br>(p.adj) |
| --- | --- | --- | --- | --- | --- | --- |
| Inferior<br>Frontal Cortex | 3.56 (0.39) | 2.41 (0.38) | 2.60 (0.18) | 0.0115<br>(**0.0173**) | 0.0031<br>(**0.0093**) | 0.5211<br>(0.5211) |
| Precentral<br>Cortex | 2.96 (0.12) | 2.09 (0.25) | 1.67 (0.45) | 0.0126<br>(**0.0378**) | 0.0303<br>(**0.0454**) | 0.2443<br>(0.2443) |
| Middle<br>Temporal Cortex | 2.60 (0.21) | 1.90 (0.15) | 1.70 (0.31) | 0.0023<br>(**0.0069**) | 0.0169<br>(**0.0254**) | 0.3974<br>(0.3974) |
| Superior<br>Parietal Cortex | 2.69 (0.21) | 2.08 (0.17) | 1.72 (0.09) | 0.0023<br>(**0.0069**) | 0.0003<br>(**0.0009**) | 0.0453<br>(**0.0453**) |
| Anterior<br>Cingulate Cortex | 2.80 (0.21) | 1.81 (0.21) | 1.45 (0.66) | 0.0022<br>(**0.0065**) | 0.0638<br>(0.0956) | 0.4427<br>(0.4427) |
| Primary<br>Visual Cortex | 2.99 (0.25) | 2.51 (0.25) | 2.33 (0.38) | 0.0640<br>(0.0960) | 0.0426<br>(0.0960) | 0.4797<br>(0.4797) |
| Putamen | 2.21 (0.19) | 1.80 (0.13) | 1.48 (0.26) | 0.0097<br>(**0.0280**) | 0.0187<br>(**0.0280**) | 0.1442<br>(0.1442) |
| Globus Pallidus | 0.41 (0.24) | 0.05 (0.03) | 0.09 (0.05) | 0.0194<br>(**0.0459**) | 0.0306<br>(**0.0459**) | 0.2744<br>(0.2744) |
| Substantia Nigra | 0.32 (0.06) | 0.09 (0.001) | 0.08 (0.01) | 0.0017<br>(**0.0026**) | 0.0011<br>(**0.0026**) | 0.2453<br>(0.2453) |
| Thalamus | 0.67 (0.15) | 0.36 (0.14) | 0.25 (0.11) | 0.0385<br>(0.0578) | 0.0056<br>(**0.0168**) | 0.3787<br>(0.3787) |
| Tegmentum<br>(Midbrain) | 0.41 (0.05) | 0.15 (0.02) | 0.13 (0.01) | <0.0001<br>(**<0.0001**) | <0.0001<br>(**<0.0001**) | 0.3294<br>(0.3294) |

Associations between ante-mortem Addenbrooke’s Cognitive Examination–Revised (ACE-R) scores and post-mortem cortical synaptic density were assessed using linear mixed-effects models adjusted for age and the proportion of time from symptom onset to death at which ACE-R was obtained. Associations between ante-mortem [^11^C]UCB-J PET and post-mortem synaptic density were evaluated by correlating regional [^11^C]UCB-J BP_ND_ with regional synaptic density across subjects and regions, allowing for repeated measures within subjects and including a covariate for the proportion of time from symptom onset to death, at the time of PET undertaken.

### PSP brain tissue shows region-specific cortical and subcortical synapse loss

We assessed synapse loss in PSP by quantifying the density of intact synapses in post-mortem brain tissue of individuals with PSP using immunofluorescence co-labelling. The PSP brain tissue was derived from study participants who underwent [^11^C]UCB-J PET during life [26,28], while the control brain tissue was obtained from neurologically healthy donors with mild age-related pathology (Table 1). Selection of 11 brain regions was based on differential PSP pathology severity [46,47] and predicted vulnerability to synaptic loss indicated by previous [^11^C] UCB-J PET studies [26,28]. Regions of interest were defined using transcortical tile scans encompassing all neocortical layers in cortical regions (Fig 1A) and anatomically delineated subregional sampling of deep grey nuclei and midbrain structures (Fig 1B). Pre- and post-synaptic terminals were immunolabeled for Bassoon and Homer1, respectively (Fig 1C), and intact synapses were defined by the proximity of the immunofluorescence signal of these markers (Fig 1D).

**Figure 1.**
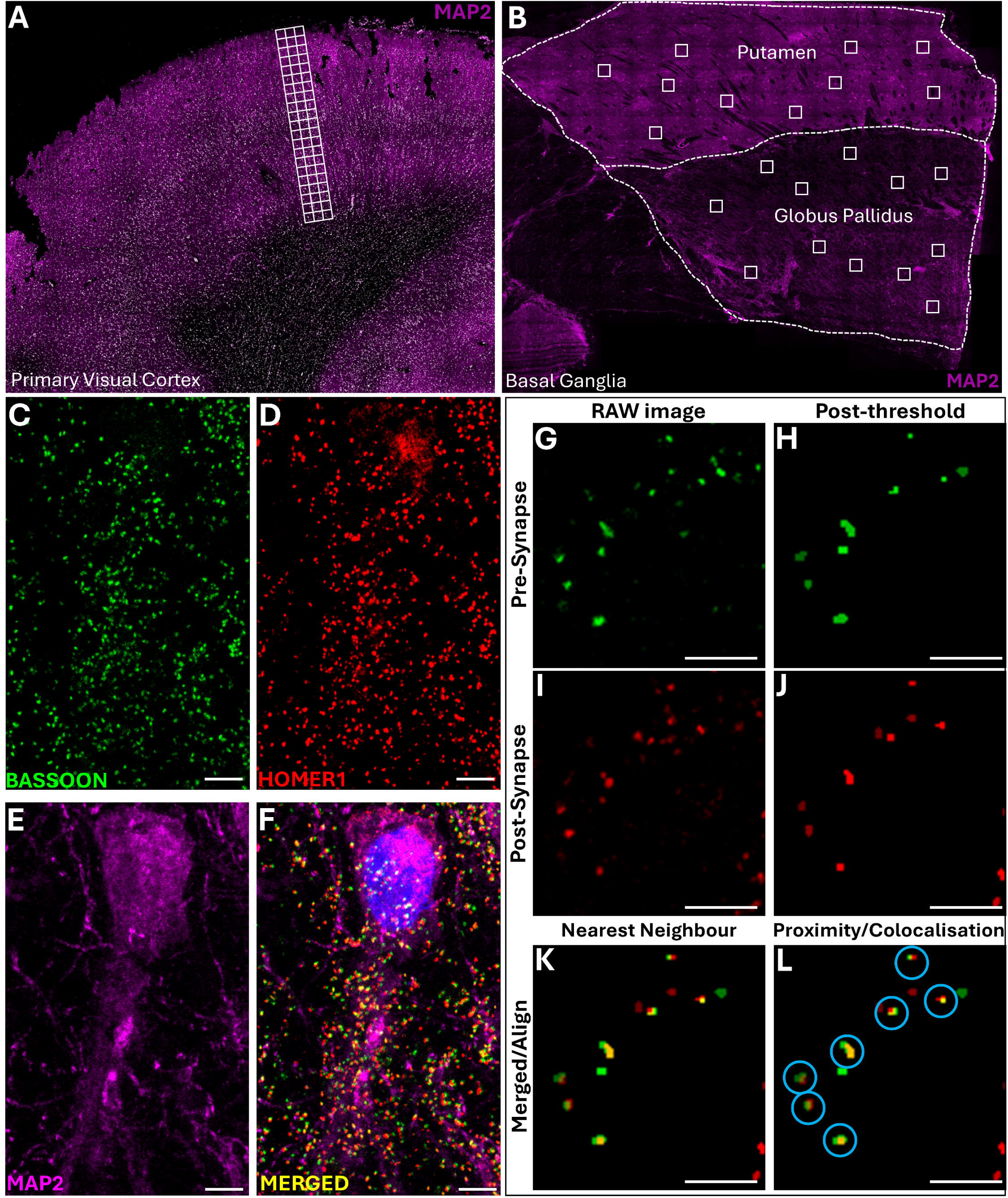
Digital pathology quantification pipeline of post-mortem synapse density. **(A)** Cortical tile scans spanning the full thickness of the cerebral cortex (reference images from control primary visual cortex). **(B)** Subcortical delineation and ROI selection. Anatomical landmarks were defined by MAP2 immunostaining, putamen and globus pallidus delineated in representative image from control basal ganglia section. **(C-F)** Representative confocal images from control inferior frontal. **(C)** Pre-synaptic marker Bassoon (green). **(D)** Post-synaptic marker Homer1 (red). **(E)** Dendritic marker MAP2 (magenta). **(F)** Merged image including DAPI nucleic stain. **(G-L)** Schematic of synapse quantification workflow. **(G-H)** Object- and intensity-based segmentation of pre-synaptic puncta. **(I-J)** Object- and intensity-based segmentation of post-synaptic puncta. **(K-L)** Nearest-neighbour analysis performed on aligned image, followed by pairing using a proximity and colocalisation algorithm. **(L)** Blue rings denote confirmed synapses. Scale bars 5um.

We showed a marked reduction in intact synapses in donors with PSP compared to controls (F(21,100) = 109.8, *p* < 0.0001) (Supplementary Table 1). The degree of synapse loss was region-specific, with the most pronounced reductions found in subcortical and brainstem areas associated with PSP pathology [46]: globus pallidus (−82.4%, *p* = 0.0245), thalamus (−65.8%, *p* = 0.0058), putamen (−25.6%, *p* = 0.0026), substantia nigra (−73.2%, *p* = 0.021) and midbrain tegmentum (−65.8%, *p* = 0.0002) (Fig 2B) (Supplementary Table 1). Sex and age at death did not influence post-mortem synaptic density (p = 0.8 and 0.7, respectively), in line with *in vivo* reports [53].

**Figure 2.**
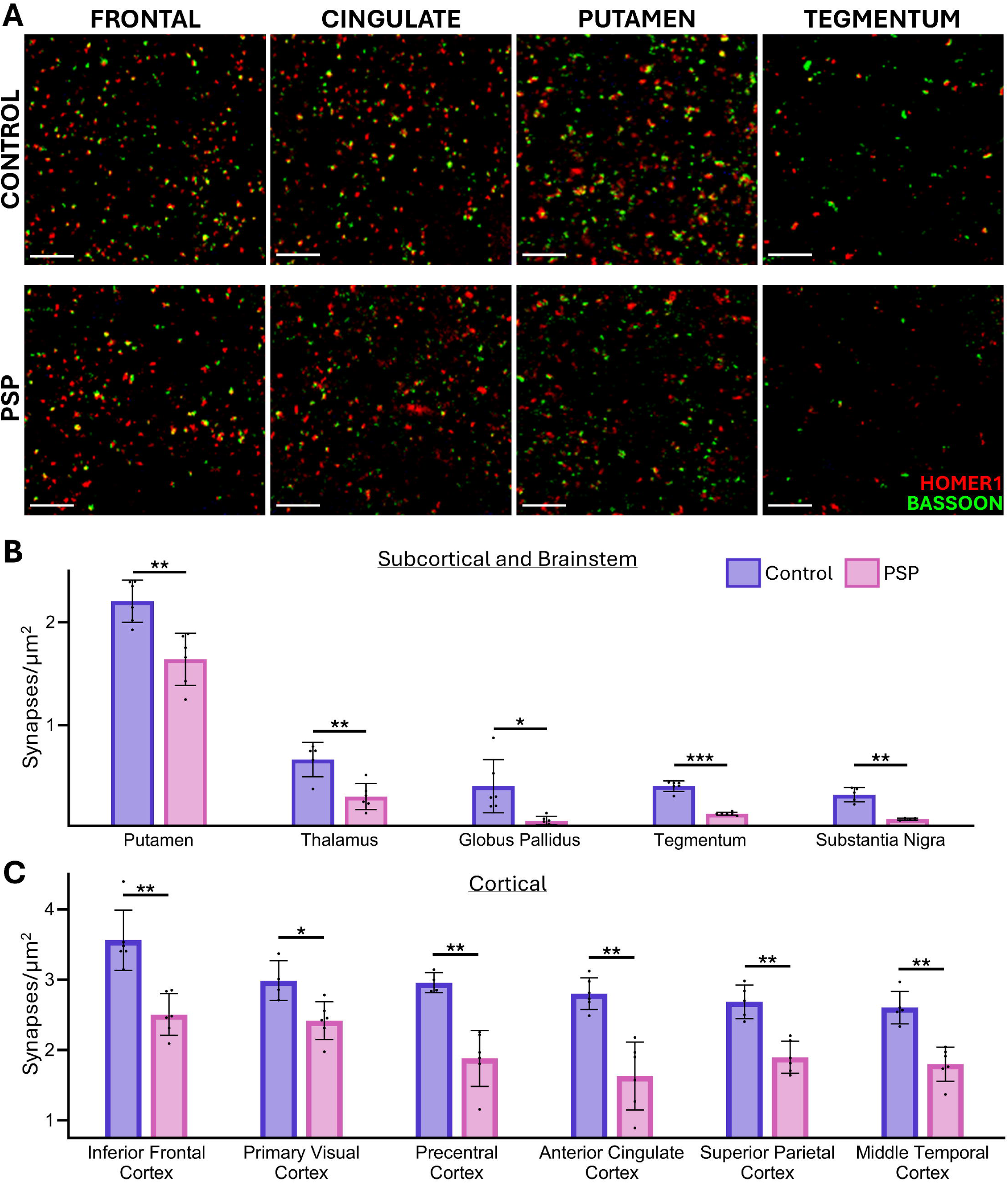
Regional synaptic density in post-mortem PSP and control brain tissue. **(A)** Representative confocal immunofluorescence images showing merged presynaptic Bassoon and post-synaptic Homer1 staining. Upper panels: control; lower panels: PSP. **(B-C)** Mean synaptic density (synapses/µm^2^ ± SD) in subcortical and brainstem **(B)** and cortical areas **(C)** of PSP and control donors. Statistical significance following false discovery rate (FDR) correction for multiple comparisons is indicated as p <0.05 *, <0.01 **, <0.001 ***.

In the neocortex, synaptic density was also significantly reduced, however, to a lesser degree (Fig 2C). The anterior cingulate cortex (−41.8% *p* = 0.0018) and precentral gyrus (−36.4%, *p* = 0.0016) exhibited the highest levels of synaptic depletion within the neocortex, highlighting these regions as particularly vulnerable to PSP pathology. Moderate synaptic loss was noted in the superior parietal lobule (−29.3%, *p* = 0.0016), inferior frontal gyrus (−29.6%, *p* = 0.0018) and middle temporal gyrus (−30.9%, *p* = 0.0016). Mild, yet significant, synaptic loss was recorded in the primary visual cortex (−19.0%, *p* = 0.0196) (Fig 2C) (Supplementary Table 1).

We next measured synaptic density across different cortical depths, delineating superficial, middle and deeper cortical regions based on tile scan coordinates (Fig 3A-B). Using a mixed effect model, we found that in both controls and PSP, the cortical synapse density is highest in the superficial versus the deeper cortical regions (F(2,152) = 48, p < 0.0001), with however no statistically significant cortical depth-specific difference between control and PSP. (Fig 3B, and Supplementary Table 2).

**Figure 3.**
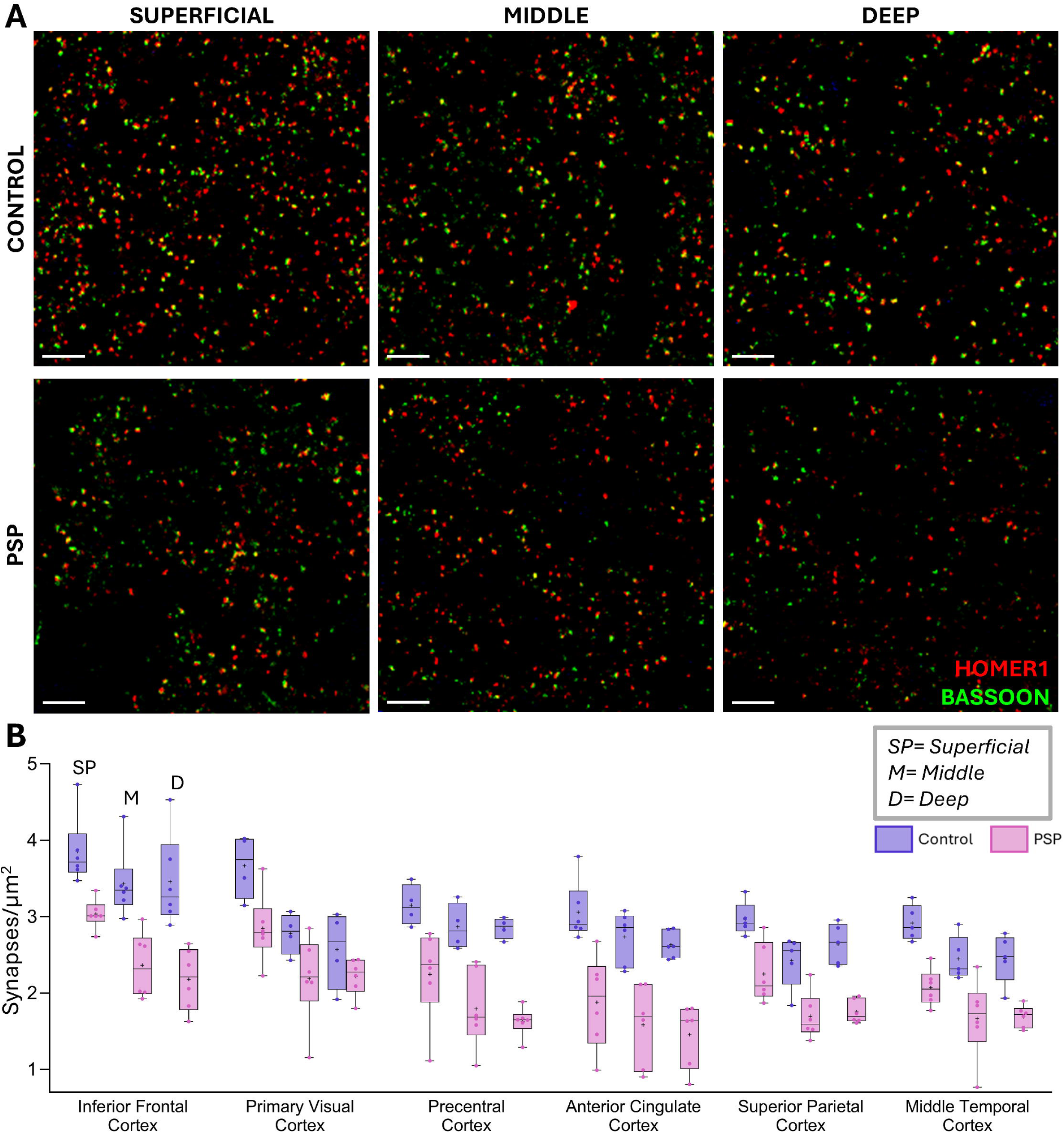
Transcortical synaptic densities in PSP and control brain tissue. **(A)** Representative confocal images of Bassoon and Homer1 in the anterior cingulate cortex (upper row:control; lower row: PSP). Representative confocal images of presynaptic (Bassoon) and postsynaptic (Homer1) staining in the anterior cingulate cortex. Upper panels: control; lower panels: PSP. Images are shown from superficial, middle, and deep cortical depth (left to right). Scale bars 5µm. **(B)** Quantification of mean synaptic density across cortical layers in control (purple) and PSP (pink) donors: superficial (S), middle (M), and deep (D). In the box plots, boxes represent the interquartile ranges, whiskers denote the minimum to maximum values, individual points represent cases, and crosses indicate the mean.

### Synaptic loss is proportional to tau pathology staging and tau burden

To assess whether disease stage influences synaptic loss, we quantified synaptic density in relation to the PSP tau pathology stages (based on the severity and distribution of the tau pathology as outlined by Kovacs et al [46]). Across most regions, synaptic loss was more pronounced in donors with advanced-stage disease (PSP tau pathology stage 5/6), compared to those at PSP tau pathology stage 4 (Fig 4B-D). Notably, early-affected regions such as the globus pallidus, substantia nigra and midbrain tegmentum exhibited comparable synaptic loss regardless of disease stage, suggesting that these structures may reach a lower limit of synaptic density by stage 4.

**Figure 4.**
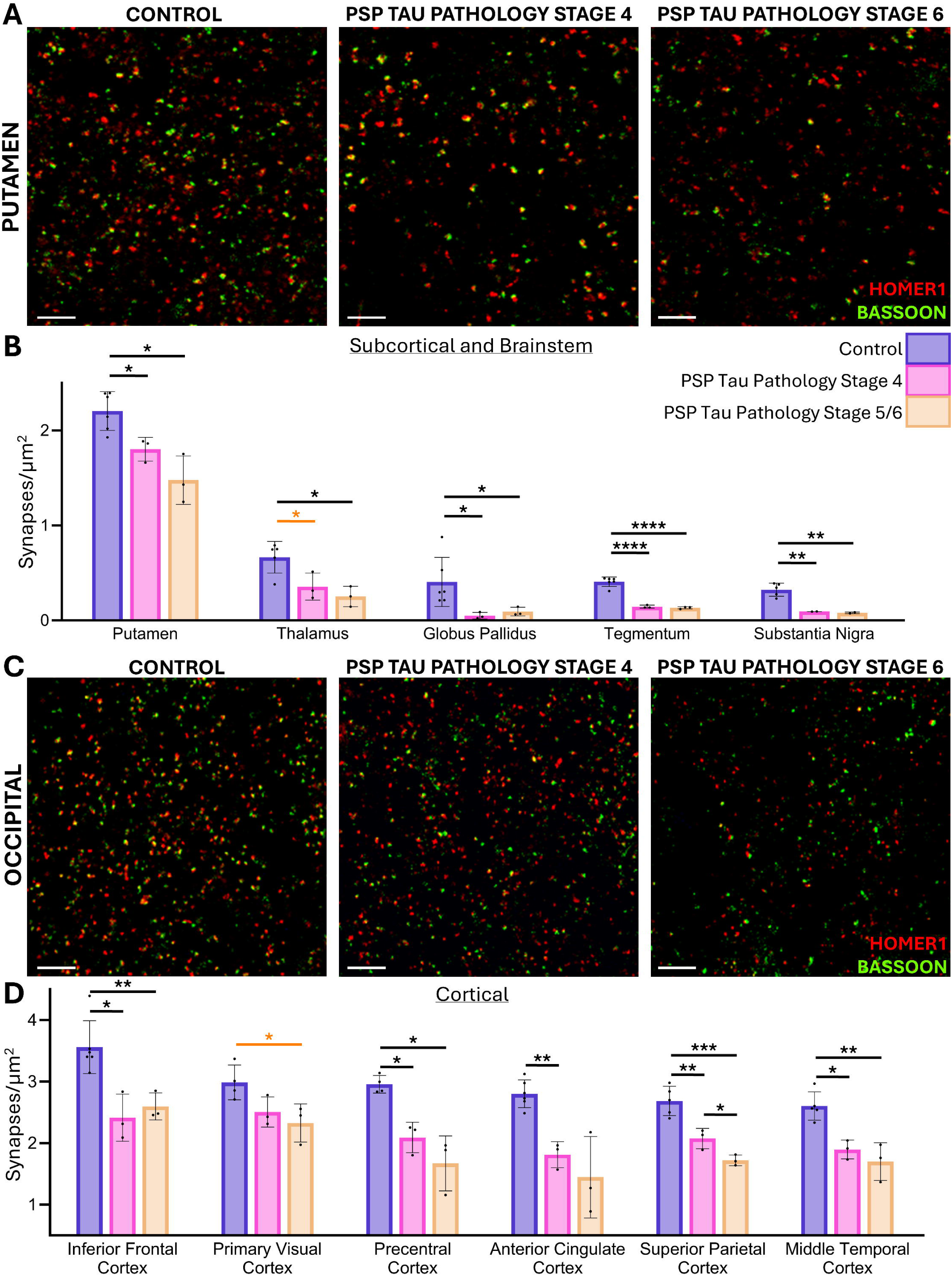
Synaptic density across PSP tau pathology stages. **(A)** Representative confocal images of presynaptic (Bassoon) and postsynaptic (Homer1) staining in putamen. **(B)** Mean synaptic density (synapses/µm² ± SD) in controls, PSP tau pathology stage 4, and PSP tau pathology stage 5/6. **(C)** Representative confocal images of the primary visual cortex in the same donors shown in **(A)**. **(D)** Mean synaptic density (synapses/µm² ± SD) in controls, PSP tau pathology stage 4, and PSP tau pathology stage 5/6. Significant differences are denoted as * p<0.05, ** p<0.01, *** p<0.001, ****p<0.0001. False discovery rate (FDR)-adjusted *p*-values for regional comparisons are shown in black; unadjusted *p*-values are shown in orange.

Using a two-way ANOVA and a mixed-effect model, the relationship between brain region and PSP tau pathology stage was tested. We showed that regional synaptic density differed across brain regions (F(2.836, 22.69) = 217.8, *p* < 0.0001), and that there was a significant effect of disease stage on synaptic density (F(2,9) = 46.19, *p <* 0.0001) demonstrating that synaptic density varied across disease stage. Furthermore, there was a significant interaction between disease stage and brain region (F(20,80) = 3.803, *p* < 0.0001) indicating that the effect of disease stage on synaptic density differed depending on the brain region examined. Post-hoc analysis revealed a significant reduction in the superior parietal lobule between stage 4 and stage 5/6 PSP cases (*p* = 0.0453) (Table 2).

Comparing PSP donors with controls, synaptic loss was significantly more severe in those at PSP tau pathology stage 5/6. This was especially evident in the thalamus (*p* = 0.0168) and the superior parietal lobule (*p* = 0.0009). The primary visual cortex approached significance at PSP tau pathology stage 5/6 (unadjusted *p* = 0.0426, adjusted *p* = 0.096) (Table 2). In contrast, stage 4 patients showed a comparatively milder proportion of synaptic reductions in these same regions. Only the superior parietal lobe reached statistical significance for direct comparisons between stage 4 and 5/6, likely due to small group sizes.

PSP tau pathology stage 4 donors also displayed significant synaptic loss across the cortex, compared to controls, particularly in the anterior cingulate cortex (*p* = 0.0065), precentral gyrus (*p* = 0.0378) and the middle temporal cortex (*p* = 0.0069). The thalamus approached statistical significance at PSP tau pathology stage 4 (unadjusted *p* = 0.0385, adjusted *p* = 0.0578).

To assess the degree to which regional tau burden influences synapse loss, we quantified tau area fraction in the same regions. Grey matter ROIs were selected in the cortex, and entire subcortical and brainstem tissue ROIs were produced for the area fractions to compare to synaptic density. Across all regions and all patients, there was a significant increase in tau pathology, on average 5.34 fold (1.22-15.57), compared to control donors (F(21, 33.13) = 13.74, p < 0.0001). In the cortex, the greatest tau burden was in the precentral gyrus (0.58%) and the inferior frontal gyrus (0.23%), with the lowest burden found in the primary visual cortex (0.07%). In the basal ganglia, the highest tau burden was found in the thalamus (0.55%), followed by the globus pallidus (0.37%) and the putamen (0.13%). In the brainstem, the substantia nigra had the highest tau burden (0.98%), followed by the tegmentum (0.61%). The difference in tau pathology between patients and controls, was only statistically significant within the thalamus after correcting for multiple comparisons (p=0.0079).

To test the relationship between tau burden and synapse loss, we examined the association between tau area fraction and synapse density. Spearman’s rank correlation demonstrated a significant negative association between tau area fraction (%) and synapses per µm² (rho = −0.522, *p* < 0.0001; Fig 5A). Simple linear regression confirmed that tau burden predicted synapse density (F(1, 47) = 14.7, *p* = 0.004), accounting for 24% of the variance (R² = 0.238).

**Figure 5.**
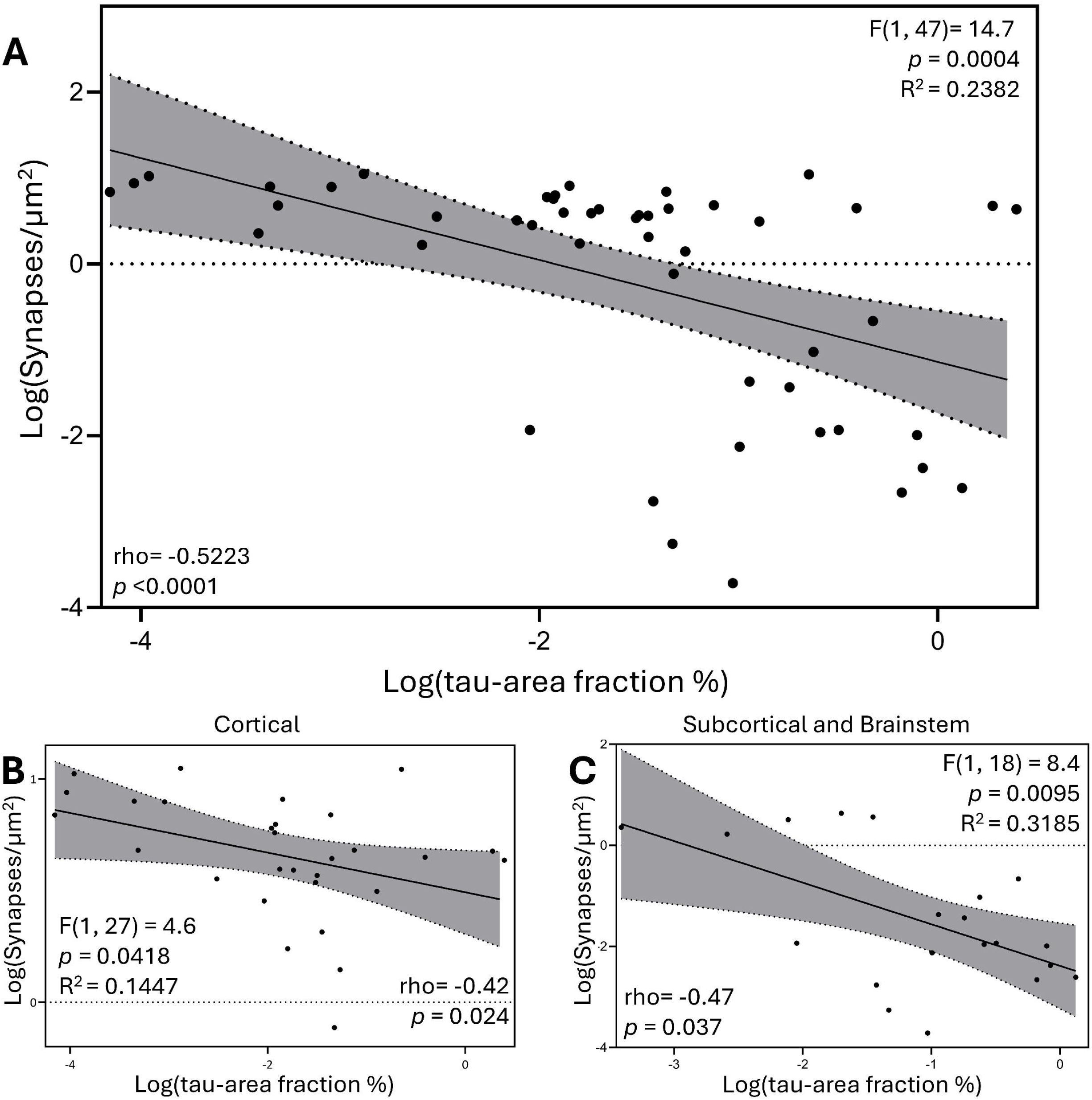
Association between synaptic density and tau burden in PSP. **(A)** Spearman’s rank correlation between log-transformed synapse density and tau area fraction with fitted linear regression and 95% confidence intervals (shaded). Reported Spearman’s rho and p-values correspond to the correlation analysis. **(B)** Correlation between cortical log-transformed synapse density and tau area fraction. **(C)** Correlation between subcortical and brainstem log-transformed synapse density and tau area fraction.

Within the cortex, tau burden was negatively associated and synapse density (rho = −0.42, *p* = 0.024), and linear regression confirmed a significant predictive effect (F(1, 27) = 4.6, *p* = 0.042), explaining 14% of the variance (R² = 0.144; Fig 5B). In subcortical and brainstem regions, a negative association was observed as well (rho = −0.47, *p* = 0.037), with linear regression demonstrating a stronger predictive relationship, F(1, 18) = 8.4, *p* = 0.0095, explaining 32% of the variance (R² = 0.318; Fig 5C). Direct comparison of regional effects revealed a significantly stronger negative association in subcortical and brainstem regions relative to cortical areas (F(1, 45) = 10.9, *p* = 0.0019).

### Post-mortem synaptic density correlates with cognitive decline and regional ante-mortem [^11^C] UCB-J PET signal

Next, we tested whether cortical post-mortem synaptic loss was associated with ante-mortem cognitive performance on the revised Addenbrooke’s Cognitive Examination (ACE-R), allowing for age and time from ACE-R assessment to death, as covariates. There was a positive correlation between post-mortem cortical synaptic density and ante-mortem cognitive performance on the ACE-R (F(3, 32) = 7, β = 11.4, standard error = 5.8, p = 0.01, adjusted R^2^ = 0.25) (Fig 6A).

**Figure 6.**
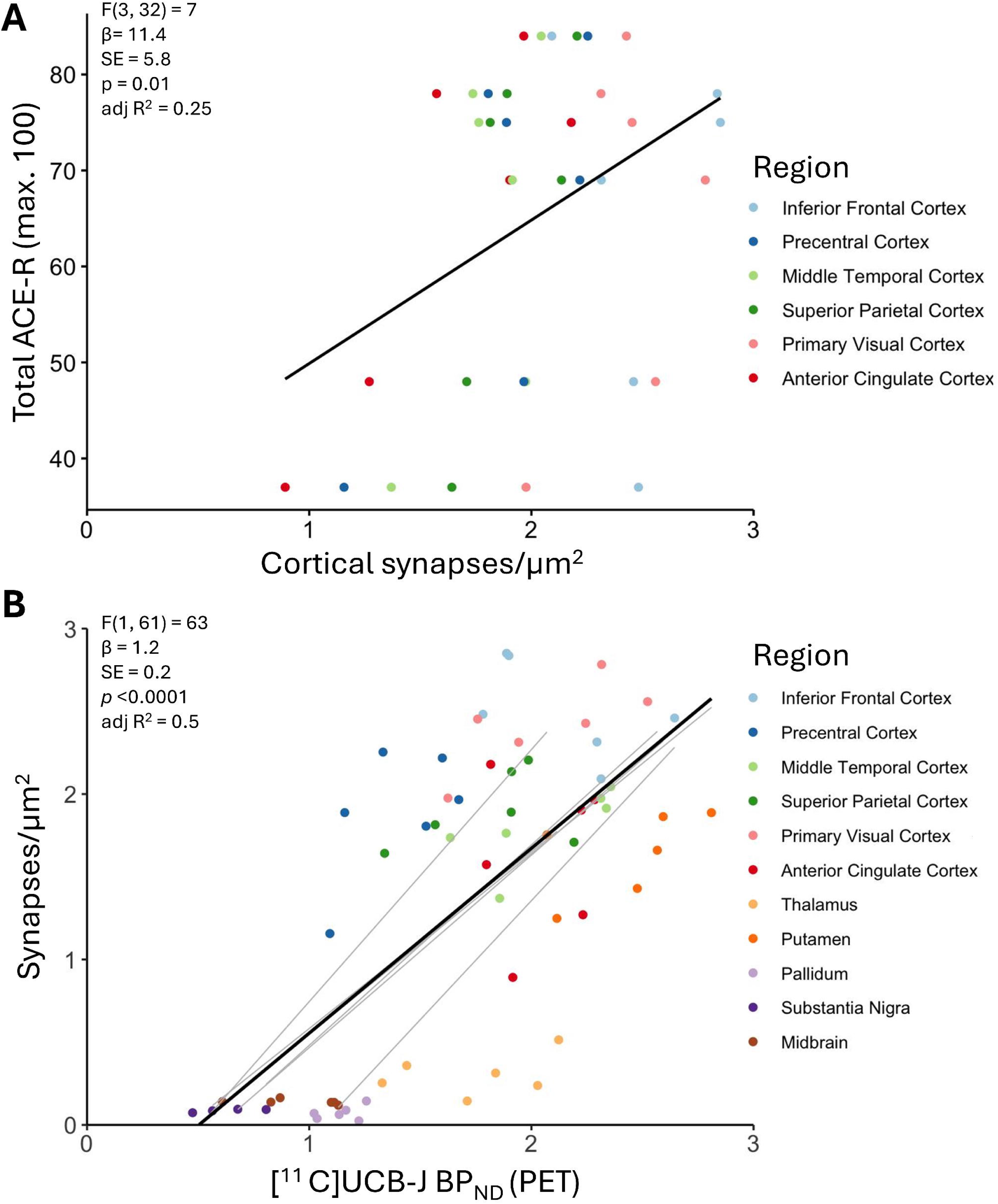
Association between ante-mortem cognitive and [^11^C]UCB-J PET measures and post-mortem synaptic density. **(A)** Correlation between ante-mortem Addenbrooke’s cognitive examination revised score and post-mortem cortical synaptic density (synapses/µm^2^). **(B)** Correlation between regional post-mortem synaptic density (synapses/µm^2^) and ante-mortem [^11^C]UCB-J non-displaceable binding potential (BP_ND_) across regions and patients. Individual patients shown as grey lines; the overall model fit shown in black.

Finally, we investigated whether *in vivo* synaptic PET imaging using [^11^C]UCB-J reflects histologically-defined synaptic density. Specifically, a linear mixed-effects model demonstrated a positive association between regional ante-mortem [^11^C]UCB-J BP_ND_ and regional post-mortem synaptic density across 11 cortical and subcortical brain regions in all six individuals with PSP (F(1, 61) = 63, β = 1.2, SE = 0.2, R² = 0.50, p < 0.0001; Fig 6B). This means that 50% of the variance in post-mortem synaptic density can be explained by ante-mortem [^11^C]UCB-J PET signal, providing strong validation of [^11^C]UCB-J as a proxy measure of synaptic density. Importantly, this relationship held after accounting for covariates including proportion of disease duration at time of PET scan, which had a comparatively small but significant negative effect on post-mortem synaptic density (F(1, 61) = 4, β = −2.2, SE = 1.0, p = 0.04). These findings support the specificity of [^11^C]UCB-J BP_ND_ to detect synaptic loss, rather than simply reflecting global disease progression.

## Discussion

There are three key findings of this study. First, by developing a bespoke histopathological imaging pipeline that identifies structurally intact synapses, we showed a marked and region-specific loss of synapses in both cortical and subcortical brain regions of individuals with PSP compared with controls. Second, we demonstrated that this loss is associated with disease severity, as measured by tau pathology burden and ante-mortem cognition. Third, by integrating ante-mortem [¹¹C]UCB-J PET with post-mortem quantification of intact synapses in the same individuals, we provide direct evidence that *in vivo* [^11^C]UCB-J PET binding potential reliably reflects synaptic density.

Our histological pipeline enabled accurate quantification of synapse preservation using co-labelling for pre- and post-synaptic markers, Bassoon and Homer1, respectively. Historically, synapse quantification typically relied on techniques measuring synaptic protein abundance (e.g. synaptophysin) [14,16,36,37,54,55]. Such approaches do not necessarily define synapse densities as these proteins are not exclusively localised in boutons [39,40]; as such, intact synapses are more accurately defined by the spatial apposition of pre- and post-synaptic terminal markers. While electron microscopy remains the gold standard of structural investigation of synapse [17,56–59]; its limited scalability precludes large multi-subject, multi-region studies. Other imaging studies have focused on individual synapse markers [60–62], few brain regions [23,61,63] or preclinical models [50,64,65]. Our pipeline overcomes these limitations by enabling: (i) distinction of intact synapses over degenerate or nascent synapses; (ii) scalability across larger cohorts and multiple brain regions; (iii) construction of large regions-of-interest; and (iv) robust and reproducible image analysis across regions and human samples.

Applying this method, we identified marked and regionally selective synaptic loss in PSP. Subcortical and brainstem structures classically implicated in PSP pathology, including the globus pallidus, thalamus, substantia nigra, and midbrain tegmentum, exhibited the most severe synaptic depletion, whereas cortical regions showed more moderate loss, with relative sparing of primary visual cortex. This distribution broadly parallels established patterns of tau pathology and disease progression in PSP [46,66], supporting the concept that synapse loss reflects region-specific vulnerability to tau-mediated neurodegeneration. Notably, early-affected regions demonstrated severe synaptic loss already by intermediate tau pathology stage, with comparatively limited additional decline at later stages. This pattern may indicate that synaptic density approaches a lower limit in these regions earlier in the disease course.

Cortical synaptic loss, although less severe than in subcortical and brainstem regions, was nonetheless marked, particularly in the precentral, frontal and parietal cortices. Neuropathological and imaging studies consistently describe prominent subcortical degeneration in PSP, especially involving the midbrain and basal ganglia, with more selective cortical involvement that most frequently affects frontal regions and motor cortex [67]. In contrast, widespread temporal or cingulate atrophy is not typically a defining feature of PSP [68]. Our observation of substantial synaptic depletion across several cortical regions, including areas not universally characterised by severe macroscopic atrophy, indicates that synaptic degeneration may exceed or precede overt neuronal loss and volume reduction in these regions [26,28]. Although neuronal density was not directly quantified, the apparent disproportion between cortical synaptic loss and reported patterns of atrophy suggests a partial dissociation between synaptic and neuronal degeneration in PSP. These findings are consistent with a model in which synapse loss constitutes a major and potentially early component of cortical involvement, even where structural MRI changes are modest.

The regional synaptic losses observed in this study align with in vivo estimates of synaptic loss in PSP from PET imaging studies [26,28], even after adjusting for progression between the time of imaging and death, and are also consistent with synapse densities reported in a single study utilising colocalised pre- and post-synaptic puncta staining in the putamen and frontal cortex of three PSP post-mortem cases [63]. The few other studies reporting synapse loss in PSP have relied on biochemical or histological quantification of synaptophysin levels. Compared with our findings, these studies report either a greater degree of cortical synapse loss [37], a lower degree of cortical loss with no overt reduction in the anterior cingulate cortex [62], or synapse loss largely restricted to the globus pallidus, with paradoxically increased cortical synaptic protein expression [21]. These differences likely reflect methodological variation, as earlier studies used presynaptic protein abundance as a proxy for synaptic density, whereas our approach quantifies structurally intact synapses defined by colocalisation of pre-and post-synaptic markers.

Synaptic loss has long been implicated in cognitive decline, as demonstrated by both in vitro and in vivo studies. Many individuals with PSP exhibit prominent behavioural and cognitive symptoms early in the disease [42,69], resembling the behavioural and language features of FTD [70,71]. In FTD, synaptophysin reduction is most marked in superficial cortical layers [37,72]. Accordingly, the effects of FTD and PSP on cortical electrophysiology (recorded by magnetoencephalogram) was best explained by individual difference in synaptic functional loss (from [^11^C]UCB-J PET) on superficial cortical layers [73]. While this study did not confirm an effect of cortical depth on disease-associated synaptic loss, this may also be a reflection of the limited sample size or the distinction between synapse loss and loss of synaptic function.

Across regions, synaptic density declined with increasing tau pathology stage and showed a significant inverse association with regional AT8-immunoreactive tau burden. These findings support in vivo imaging studies that have demonstrated spatial coupling between tau accumulation and synaptic loss in PSP [27], and AD [74]. However, fibrillar tau burden accounted for only a proportion of the observed variance in synaptic density, indicating that aggregated tau alone does not fully explain synaptic degeneration. AT8 immunohistochemistry primarily detects hyperphosphorylated, fibrillar tau species that may represent a downstream marker of pathology. Earlier synaptotoxic mechanisms, including effects of soluble or oligomeric tau species not captured by conventional histological staining [75], may contribute substantially to synaptic vulnerability [5,6,8,12,74]. While synapse degeneration may precede tau aggregation in vulnerable regions, additional processes such as neuroinflammation, glial dysfunction, or axonal disconnection may further modulate regional synapse loss.

The clinical relevance of synaptic loss was underscored by the positive association between post-mortem cortical synaptic density and ante-mortem cognitive performance on the ACE-R. This finding provides direct post-mortem confirmation that synaptic integrity is a critical pathological substrate of cognitive impairment in PSP, consistent with seminal observations in AD [16,18,59]. These data reinforce the concept that synapse loss, rather than neuronal loss alone, underlies cognitive decline in tau-mediated neurodegeneration.

*In vivo* [^11^C]UCB-J PET imaging has been applied across multiple neurodegenerative diseases, yet the extent to which the PET signal reflects true synaptic density has remained uncertain. In this first-of-its-kind [^11^C]UCB-J PET to post-mortem study we show a strong correlation between ante-mortem [¹¹C]UCB-J binding potential and post-mortem synaptic density across individuals and anatomically matched brain regions, even after accounting for disease duration at the time of imaging. By bridging in vivo imaging with structural synapse quantification, our findings extend prior autoradiographic work and establish [¹¹C]UCB-J PET as a robust surrogate marker of synaptic density in clinical studies.

The following limitations should be acknowledged. First, the cohort size was small, reflecting the rarity of individuals who undergo ante-mortem [¹¹C]UCB-J PET imaging and subsequently donate brain tissue. Replication across larger, independent cohorts encompassing a broader range of PSP phenotypes would be desirable to confirm generalisability. Second, cortical depth analysis was based on a three-layer model rather than lamina-specific markers. Future studies incorporating layer-resolved markers will enable more precise dissection of circuit-level vulnerability in PSP.

In conclusion, we demonstrate widespread region-dependent synapse loss in PSP that correlates with tau pathology, cognitive impairment, and ante-mortem [¹¹C]UCB-J PET binding potential. These findings provide direct histopathological validation of synaptic PET imaging and establish synapse loss as a key intermediate phenotype linking molecular pathology to clinical decline. The ability to accurately quantify synaptic loss *in vivo* offers a powerful platform for mechanistic studies and therapeutic trials targeting synaptic preservation in multiple neurodegenerative disorders.

## Data Availability

Anonymized post-mortem and PET data used for this analysis are available on request. Further participant-specific information, images or samples can be requested but are likely to require a data/material transfer agreement to adhere to consent restrictions including protection of confidentiality.

## Notes

### Competing Interest Statement

The authors have declared no competing interest.

### Author Declarations

All tissue was obtained from the Cambridge Brain Bank under the Neuropathology Research in Dementia committee(Research Ethics Committee reference 16/WA/0240), which gave full ethical approval for this work.

### Funding

- Wellcome Trust: 220258
- Alzheimer’s Research UK: 2024-006
- Alzheimer's Research UK Eastern Network, the Cambridge Centre for Parkinsons Plus: RG95450
- NIHR Cambridge Biomedical Research Centre: NIHR203312
- Alzheimer's Research UK, Race Against Dementia: ARUK-RADF2021A-010
- Medical Research Council: MR/X006867/1

