## Supplementary Tables 1 and 2 for "Synapse loss in Progressive Supranuclear Palsy *post-mortem* reflects clinical and pathological disease severity and ^11^C-UCB-J PET *in vivo*"

### Supplementary material

| Brain region | Control synapses/µm$^2$ (SD) | PSP synapses/µm$^2$ (SD) | Proportional density loss % (SD) | *p value* | *adj. q value* |
|---|---|---|---|---|---|
| Inferior Frontal Cortex | 3.56 (0.39) | 2.51 (0.27) | -29.62 (7.60) | 0.0008 | 0.0018 |
| Primary Visual Cortex | 2.99 (0.25) | 2.42 (0.25) | -19.04 (8.21) | 0.0178 | 0.0196 |
| Middle Temporal Cortex | 2.60 (0.21) | 1.80 (0.22) | -30.86 (8.50) | 0.0003 | 0.0016 |
| Precentral Cortex | 2.96 (0.12) | 1.88 (0.36) | -36.40 (12.27) | 0.0006 | 0.0016 |
| Anterior Cingulate Cortex | 2.80 (0.21) | 1.63 (0.44) | -41.82 (15.73) | 0.0010 | 0.0018 |
| Superior Parietal Cortex | 2.69 (0.21) | 1.90 (0.21) | -29.29 (7.74) | 0.0004 | 0.0016 |
| Putamen | 2.21 (0.19) | 1.64 (0.23) | -25.61 (10.51) | 0.0019 | 0.0026 |
| Thalamus | 0.67 (0.15) | 0.30 (0.12) | -54.38 (17.29) | 0.0047 | 0.0058 |
| Globus Pallidus | 0.41 (0.24) | 0.07 (0.04) | -82.36 (9.57) | 0.0245 | 0.0245 |
| Substantia Nigra | 0.32 (0.06) | 0.09 (0.01) | -73.20 (2.58) | 0.0013 | 0.0021 |
| Tegmentum | 0.41 (0.05) | 0.14 (0.01) | -65.76 (3.24) | $<0.0001$ | 0.0002 |

**Supplementary Table 1. Summary statistics of synaptic density in PSP and controls**. Group-wise mean (standard deviation) synaptic density is shown for PSP and control donors, together with proportional synaptic loss in PSP relative to controls (standard deviation). P-values are derived from post hoc pairwise comparisons following one-way ANOVA with Welch's correction for unequal variances. Q-values indicate false discovery rate (FDR)-adjusted P-values to account for multiple comparisons.

| Cortical area | Cortical Region | Control synapses/µm² (SD) | PSP synapses/µm² (SD) | % Loss (SD) | Adj. *p* val | Cohen's D |
|---|---|---|---|---|---|---|
| Superficial cortical area | Inferior Frontal Cortex | 3.85 (0.45) | 3.03 (0.20) | -21.29 (5.05) | **0.01** | **-2.4** |
| | Precentral Cortex | 3.15 (0.27) | 2.24 (0.61) | -28.7 (17.53) | **0.02** | **-1.8** |
| | Middle Temporal Cortex | 2.92 (0.23) | 2.07 (0.24) | -28.99 (8.26) | **0.02** | **-3.6** |
| | Superior Parietal Cortex | 2.97 (0.22) | 2.25 (0.39) | -24.18 (12.02) | 0.05 | -2.2 |
| | Anterior Cingulate Cortex | 3.06 (0.39) | 1.88 (0.61) | -38.53 (19.91) | **0.002** | **-2.3** |
| | Primary Visual Cortex | 3.67 (0.42) | 2.85 (0.453) | -22.30 (12.35) | **0.002** | **-1.9** |
| Middle cortical area | Inferior Frontal Cortex | 3.43 (0.46) | 2.36 (0.43) | -31.17 (12.60) | **0.002** | **-2.4** |
| | Precentral Cortex | 2.87 (0.30) | 1.79 (0.51) | -37.47 (16.35) | **0.002** | **-2.4** |
| | Middle Temporal Cortex | 2.45 (0.29) | 1.67 (0.52) | -31.92 (31.28) | **0.02** | **-1.8** |
| | Superior Parietal Cortex | 2.42 (0.35) | 1.69 (0.31) | -30.15 (11.55) | **0.05** | **-2.3** |
| | Anterior Cingulate Cortex | 2.73 (0.34) | 1.58 (0.53) | -42.10 (19.42) | **0.002** | **-2.6** |
| | Primary Visual Cortex | 2.78 (0.27) | 2.19 (0.58) | -21.23 (20.72) | **0.02** | **-1.2** |
| Deep cortical area | Inferior Frontal Cortex | 3.46 (0.60) | 2.18 (0.41) | -37.0 (11.80) | **0.002** | **-2.5** |
| | Precentral Cortex | 2.85 (0.14) | 1.63 (0.19) | -42.98 (6.13) | **0.002** | **-7.1** |
| | Middle Temporal Cortex | 2.45 (0.33) | 1.69 (0.14) | -31.07 (8.42) | **0.002** | **-3.1** |
| | Superior Parietal Cortex | 2.64 (0.27) | 1.75 (0.15) | -33.66 (5.32) | **0.01** | **-4.2** |
| | Anterior Cingulate Cortex | 2.63 (0.18) | 1.46 (0.42) | -44.71 (15.79) | **0.002** | **-3.7** |
| | Primary Visual Cortex | 2.57 (0.51) | 2.22 (0.24) | -13.77 (9.45) | ns | -1.0 |

**Supplementary Table 2. Cortical synaptic density by depth and region in control and PSP donors.** Mean synaptic density (synapses/µm², standard deviation) and proportional loss relative to controls (%, standard deviation) are shown for each cortical depth and region, stratified by group (Control, PSP). P-values are derived from post-hoc contrasts of a linear mixed-effects model testing region × depth interactions between groups, with Bonferroni correction for multiple comparisons. Effect sizes for group differences are reported as Cohen's d.
